# Christianson Syndrome across the Lifespan: An International Longitudinal Study in Children, Adolescents, and Adults

**DOI:** 10.1101/2023.11.11.23298218

**Authors:** Brian C. Kavanaugh, Jennifer Elacio, Carrie R. Best, Danielle G. St. Pierre, Matthew F. Pescosolido, Qing Ouyang, Paul Caruso, Karen Buch, John Biedermann, Rebecca S. Bradley, Judy S. Liu, Richard N. Jones, Eric M. Morrow

**Author notes:** To whom correspondence should be addressed: Eric M. Morrow MD PhD, Brown University, Lab for Molecular Medicine, 70 Ship Street, Providence, RI 02912.

## Abstract

Mutations in the X-linked endosomal Na+/H+ Exchanger 6 (NHE6) causes Christianson Syndrome (CS). In the largest study to date, we examine genetic diversity and clinical progression, including cerebellar degeneration, in CS into adulthood. Data were collected as part of the International Christianson Syndrome and NHE6 (SLC9A6) Gene Network Study. Forty-four individuals with 31 unique NHE6 mutations, age 2 to 32 years, were followed prospectively, herein reporting baseline, 1-year follow-up, and retrospective natural history. We present data on the CS phenotype with regard to physical growth, adaptive and motor regression, and across the lifespan, including information on mortality. Longitudinal data on body weight and height were examined using a linear mixed model: the rate of growth across development was slow and resulted in prominently decreased age-normed height and weight by adulthood. Adaptive functioning was longitudinally examined: a majority of adult (18+ years) participants lost gross and fine motor skills over a 1-year follow-up. Previously defined core diagnostic criteria for CS (present in >85%) – namely nonverbal status, intellectual disability, epilepsy, postnatal microcephaly, ataxia, hyperkinesia – were universally present in age 6 to 16; however, an additional core feature of high pain tolerance was added (present in 91%), and furthermore, evolution of symptoms were noted across the lifespan, such that postnatal microcephaly, ataxia and high pain threshold were often not apparent prior to age 6, and hyperkinesis decreased after age 16. While neurologic exams were consistent with cerebellar dysfunction, importantly, a majority of individuals (>50% older than 10) also had corticospinal tract abnormalities. Three participants died during the period of the study. In this large and longitudinal study of CS, we begin to define the trajectory of symptoms and the adult phenotype, thereby identifying critical targets for treatment.

## INTRODUCTION

Christianson syndrome (CS) in males results from loss-of-function (LOF) mutations in the X-linked gene *SLC9A6* which encodes the endosomal Na+/H+ Exchanger 6 (NHE6).^1–4^ CS is one of the most common forms of X-linked intellectual disability.^5^ The NHE6 protein modulates intra-endosomal pH and endosomal maturation.^6,7^ Studies of the pathophysiology of CS in animals and cells lacking NHE6 have identified both neurodevelopmental and neurodegenerative features.^6,8–11^ In male patients, CS presents early with global developmental delay, lack of language, postnatal microcephaly, ataxia, epilepsy, and Angelman Syndrome-like and/or autistic features.^1^ However, little systematic data exists which establishes the life course of symptoms in people with CS into adulthood. In addition to abnormal brain development, progressive neurodegenerative and medical processes unfold with age.^1,12^ Accordingly, CS has been conceptualized as a mixed neurodevelopmental and neurodegenerative disorder.^10^

In the initial large pedigree, Dr. Arnold Christianson and colleagues reported that early neurodevelopmental abnormalities were followed by a “slow regression” starting in the second decade of life.^3^ In our prior report of 14 CS probands, we identified a history of regression in 50% of participants.^1^ Also, CS has been described to involve a progressively worsening ataxia, which appears to be associated with the high prevalence of cerebellar degeneration in patients (atrophy in 30-60% of cases).^1–4,13^ Further, human neuropathologic studies^3,12^ and NHE6-null mouse and rat models have identified cerebellar Purkinje cell loss, and as well as cortical tissue loss, with tau deposition.^8–10^ Dr. Christianson’s original description of an extended South African pedigree suggested progressive morbidity, thin body habitus, and a potential risk of early mortality^3^; however, to date the risk of early mortality has not been corroborated in larger studies.

A range of different NHE6 mutations have been reported in CS. To date, the majority of published NHE6 mutations are nonsense or splice mutations that are thought to result in complete loss-of-function of the NHE6 protein.^1–4^ Nonsense mutations appear to lead to nonsense mediated mRNA decay.^14^ Other mutation types (for example missense) have also been reported but are less common, yet may be informative to protein function.^1,15^ Mutations in CS males generally cluster in the Na^+^/H^+^ exchanger region and are less commonly found in the C-terminus cytoplasmic tail.^1,2^ These cytoplasmic tail mutations, again particularly when missense, could be highly informative given that the function of this part of the protein is not well understood. While previously reported as inherited in pedigrees, *de novo* mutations now appear to be a relatively common form of transmission.^1^ Finally, Pescosolido et al.^1^ found little genotype-phenotype correlations between specific mutations, although the sample size was limiting for this assessment.

Based on the International Christianson Syndrome and NHE6 (SLC9A6) Gene Network Study, the current study examined the largest sample to date of CS probands across the lifespan, importantly including longitudinal follow-up data and a large proportion of adults. In total this study includes 44 probands, including 11 adults. This group of patients is more than three times as large as the largest prior report^1^, with a wider age range of participants (age 2 to 32 years). Participants were enrolled as a result of a collaboration of international participants, clinicians, researchers and the Christianson Syndrome Association (CSA). With a focus on potential target symptoms for future clinical trials, the four primary aims of this study are to 1) define new mutations and the range of NHE6 mutations in CS; 2) cross-sectionally and longitudinally examine the clinical phenotype across the lifespan, as well as the adult phenotype and potential early mortality; and 3) longitudinally examine growth and maintenance of healthy weight of the probands across the lifespan. Importantly, we present new mutations, including informative missense mutations. Also, through annual follow-up, we define a prominent component of the progressive phenotype in CS involving the longitudinal attenuation of weight gain and growth, as well as loss of motor function with age. We have also observed deaths of participants. In summary, given the size and the longitudinal design, the current study permits a quantitative investigation of the CS phenotype across the lifespan, and importantly, the definition of the CS phenotype in adulthood.

## METHODS

### Procedures

This Christianson Syndrome and NHE6 (*SLC9A6*) Gene Network Study was approved by the Institutional Review Board (IRB) at Lifespan Healthcare and Brown University. Informed consent was given by the parent/guardian of the participant. The procedures involved the initial enrollment and yearly follow-ups. We generated natural history data through both retrospective medical record review and history taking, as well as annual prospective follow-up assessments. At the time of this report, one-year, prospective follow-up data on 22 patients were available.

### Recruitment and Enrollment

Enrollment criteria were adapted from Pescosolido et al.^1^ and included males at any age wherein: 1) there was a prior clinical genetic diagnosis involving mutation in the *SLC9A6* gene; 2) the participating child had an active clinician; and 3) the clinical phenotype was consistent with CS diagnostic criteria. CS probands and their extended families were enrolled. Participants were recruited through close collaboration with the family association, the Christianson Syndrome Association (CSA), participation in the biannual international conferences for CS, and via genetic diagnostic labs. Participants were predominantly enrolled remotely in this study due to our international recruitment that spans 11 countries. Materials were translated into the native language of the family and remote assessments were conducted with a translator when possible. DNA was isolated from blood or saliva samples. All *NHE6* mutations were verified by Sanger sequencing by sequencing all coding exons (1-16) and exon/intron junctions as previously described^1^ except where indicated in Table 1.

**Table 1.** NHE6 mutations found in 44 affected males across 37 families. All transcript positions are based on NM_001042537.1 and protein sequences are based on NP_001036002.1. Coordinates reported are hg19. For mutations found in more than one individual, the number of individuals is denoted in brackets under Variant Type. For mutations found in more than one family, the number of families is denoted in brackets under Inheritance. ^a^ Indicates mutations not confirmed by Sanger sequencing and data based solely on medical records.

| Mutation | cDNA Position | Protein Position | Exon/Intron | Variant Type | Inheritance | Genomic Coordinates |
| --- | --- | --- | --- | --- | --- | --- |
| 1 | 2073bp deletion including exon 1 and some of intron 1 | - | Exon 1/Intron 1 | Deletion (2) | Unknown | chrX:135066366-135068439 |
| 2 | c.1A>G | p.M1? | Exon 1 | Start Lost | Inherited | chrX:135067662 |
| 3 | c.205C>T | p.Q69* | Exon 1 | Nonsense (2) | Inherited | chrX:135067866 |
| 4 | c.335_336delTG | p.V112Gfs*13 | Exon 2 | Frameshift (2) | Inherited | chrX:135076954-135076955 |
| 5 | c.526-1G>A | - | Intron 2 | Splice | De novo | chrX:135080266 |
| 6 | c.540_547dup | p.F183fs*1 | Exon 3 | Frameshift | Unknown | chrX:135080281-135080288 |
| 7<sup>a</sup> | c.603+1G>A | - | Intron 3 | Splice | Inherited | chrX:135080345 |
| 8 | c.604-1G>T | - | Intron 3 | Splice | Inherited | chrX:135080640 |
| 9 | c.604-1G>A | - | Intron 3 | Splice (2) | Inherited | chrX:135080640 |
| 10 | c.630_631insT | p.I211Yfs*53 | Exon 4 | Frameshift (3) | Inherited | chrX:135080668 |
| 11 | c.680+1G>A | - | Intron 4 | Splice | De novo | chrX:135080718 |
| 12 | c.680+5G>A | - | Intron 4 | Splice | De novo | chrX:135080722 |
| 13 | c.899+1G>A | - | Intron 6 | Splice | Inherited | chrX:135084373 |
| 14 | c.899+1delGTAA | - | Intron 6 | Splice (2) | Unknown (1)<br>Inherited (1) | chrX:135084373-135084376 |
| 15 | c.953_954delCA | p.T318Sfs*59 | Exon 7 | Frameshift | Inherited | chrX:135092654-135092655 |
| 16 | c.1024G>A | p.G342R | Exon 7 | Missense (2) | De novo (2) | chrX:135092725 |
| 17 | c.1148G>A | p.G383D | Exon 9 | Missense (2) | De novo (2) | chrX:135095508 |
| 18 | c.1222_1226delCATAG | p.H408Nfs*2 | Exon 9 | Frameshift | De novo | chrX:135095582-135095586 |
| 19 | 120.7kb deletion from intron 10 through 3'-untranslated region | - | Intron 10 to Exon 16 | Deletion | Inherited | chrX:135098247-135218928 |
| 20 | c.1380_1389delinsGTCTTGGGAAGAC T | p.N460Kfs*12 | Exon 11 | Frameshift | Inherited | chrX:135104774-135104780 |
| 21<sup>a</sup> | c.1402_1405del | p.L468Ifs*15 | Exon 11 | Frameshift | Unknown | chrX:135104796-135104799 |
| 22 | c.1414insA | p.R472Kfs*4 | Exon 11 | Frameshift | De novo | chrX:135104808 |
| 23 | c.1462+1G>C | - | Intron 11 | Splice | Unknown | chrX:135104857 |
| 24 | c.1472G>A | p.G491D | Exon 12 | Missense | Inherited | chrX:135106498 |
| 25 | c.1498C>T | p.R500* | Exon 12 | Nonsense (3) | Inherited (2)<br>Unknown (1) | chrX:135106524 |
| 26 | c.1568G>A | p.W523* | Exon 12 | Nonsense | De novo | chrX:135106594 |
| 27 | c.1569G>A | p.W523* | Exon 12 | Nonsense | Unknown | chrX:135106595 |
| 28<sup>a</sup> | c.1631C>G | p.S544* | Exon 13 | Nonsense | De novo | chrX:135112305 |
| 29 | c.1639G>T | p.E547* | Exon 13 | Nonsense (2) | Inherited | chrX:135112313 |
| 30 | c.1710G>A | p.W570* | Exon 14 | Nonsense (2) | De novo (1)<br>Unknown (1) | chrX:135115635 |
| 31 | c.1745T>C | p.L582P | Exon 15 | Missense (1) | De novo | chrX:135122252 |

Note. All transcript positions are based on NM\_001042537.1 and protein sequences are based on NP\_001036002.1. Coordinates reported are hg19. Under Variant Type, if the mutation is found in more than one individual, the total number of individuals are indicated in brackets. Under Inheritance, if the mutation is found in more than one family, the number of families are indicated in brackets. <sup>a</sup> Indicates mutations wherein confirmation by Sanger sequencing was not performed, and the data are based on the medical record alone.

### Clinical Assessment and Medical Record Review

Clinical data were collected by telephone, video-conferencing and/or in person assessments and through medical record collection. Standardized research assessments were also filled out by parents/guardians. The Vineland Adaptive Behavior Scales-Second Edition (VABS-2)^16^ assessed adaptive functioning across the following domains: communication, social, daily living, motor, and maladaptive behavior. Irritability and hyperactivity were assessed via the Aberrant Behavior Checklist (ABC)^17^, a parent questionnaire to assess behavioral problems in individuals with developmental disorders. A detailed medical and family history questionnaire, including items specific to CS, was administered to parents/guardians. Height and weight were obtained from available medical notes. In-person standard neurological examinations included assessments of cranial nerve, motor (e.g. muscle tone and reflexes), coordination (e.g. gait), and sensory function that were performed by a board-certified neurologist (JSL). Prospective follow-up involved an annual telephone or video interview to collect information regarding changes to medical, neurological, or behavioral symptoms experienced by the proband since the family’s last contact with the study. VABS-2 and ABC were administered, and new medical records were collected.

### Statistical Analyses

The sample (n = 44) was divided into four groups, broadly based on developmental stage at enrollment: toddlers (0-5 years; n = 11), children (6-11 years; n = 9), adolescents (12-16 years; n = 13), and adults (17-32 years; n = 11). ANOVA and chi-squared analysis were used to examine group differences in clinical characteristics across developmental stages, inheritance/*de novo*, and mutation type. A subset of the group (n = 22) completed the VABS-2 at baseline and re-assessment at the study 1-year follow-up. VABS-2 subdomains were used to measure expressive language (Communication scale), interpersonal skills (Social scale), fine motor skills (Motor scale), and gross motor skills (Motor scale) were examined. The change in raw skills between baseline and follow-up was calculated to examine potential intra-individual decline and analyzed using Pearson’s correlation. Raw measurements of height/weight were converted to age-normed percentiles (WHO growth standards for 0-2 years^18^; CDC growth curves for 2-20 years^19^) and to z-scores. To model physical stature across development, we used linear mixed model (with random slopes and intercepts) for each variable of interest (i.e., height, weight) with the age of the child as the time basis. The intercept and age at time point were modeled as fixed and random effects.

## RESULTS

### CS-associated Mutations

In total, 37 distinct families were enrolled with 44 affected males. Thirty-one unique mutations were identified in the cohort (Table 1; Figure 1). Of the 31 unique mutations, there were: 15 frameshift and/or nonsense mutations; four deleterious missense mutations; nine splice mutations; two copy number variant deletions; and one mutation that removes the first methionine residue. Of those with known inheritance (29 of 37 families), 16 cases were inherited (55%) and 13 cases were *de novo* (45%). Notably, five mutations were recurrent (i.e. occurred in more than one family). Each of the following mutations occurred in two distinct families: c.899+1delGTAA (mutation 14), c.1024G>A (mutation 16), c.1148G>A (mutation 17), and c.1710G>A (mutation 30). Mutation 25, c.1498C>T, was found in three families. These recurrent mutations were inherited in three families not known to be related and were *de novo* in five families. Inheritance was unknown in the three remaining families with recurrent mutations. No strong genotype-phenotype associations were evident in the data (Supplemental Table 1).

**Figure 1.**
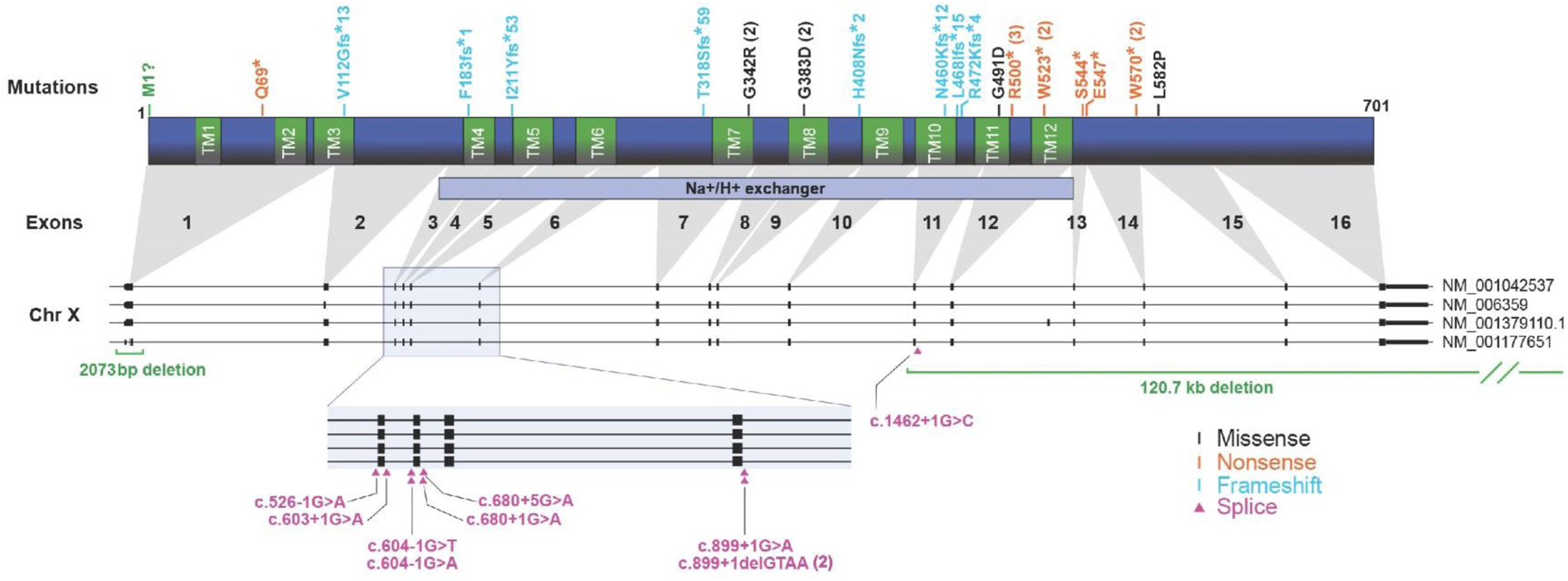
Christianson syndrome-associated NHE6 mutations. A total of 31 unique NHE6 mutations were identified in 44 CS male probands across 37 families. Pathogenic *NHE6* mutations include: frameshift and/or nonsense (n = 15, cyan or orange), deleterious missense (n = 4, black), splice (n = 9), CNVs (n = 2, green) and one that removes the first methionine residue. Out of CS pedigrees with known inheritance (78%, 29 out of 37 families), 55% were inherited (16 out of 29 pedigrees) and 45% were *de novo* (13 out of 29 pedigrees). A total of five mutations were recurrent across multiple pedigrees: c.899+1delGTAA (mutation 14, 2 pedigrees), c.1024G>A (mutation 16, 2 pedigrees), c.1148G>A (mutation 17, 2 pedigrees), c.1498C>T (mutation 25, 3 pedigrees) and c.1710G>A (mutation 30, 2 pedigrees). The following mRNA isoforms are used: NM_001042537, NM_006359, NM_001379110.1 and NM_001177651. TM=transmembrane.

### CS Core and Associated Symptoms Across the Lifespan

In total, 44 males were evaluated with *SLC9A6* mutations, ranging from age 2 to 32 years. In order to consider CS symptoms across developmental stages, cross-sectional analysis was conducted across four age groups (Table 2): 0-5 years at enrollment (n = 11); 6-11 years (n = 9); 12-16 years (n = 13); and 17-32 years (n = 11). In terms of race and ethnicity: 11% of the sample identified as Hispanic/Latino (n = 5); 89% identified as White (n = 39); 5% identified as Multi-racial (n = 2); 2% identified as Black (n = 1); and 2% identified as Other (n = 1). Participants were born in eleven different countries and seven different primary languages were represented in these families.

**Table 2.** Clinical characteristics of CS males across developmental cohorts. + Denotes numbers approaching statistical significance (p < .05). Italicized domains represent proposed core CS symptoms. Adaptive functioning score from VABS-2 is a composite of the communication, daily living skills, socialization and motor skills domain scores. GERD=Gastroesophageal reflux disease. ABC=Aberrant Behavior Checklist. VABS-2=Vineland Adaptive Behavior Scales-2^nd^ Edition.

|  | n | Total sample<br>(n = 44) | Toddlers<br>(2.5-5 yrs; n = 11) | Children<br>(6-11 yrs; n = 9) | Adolescents<br>(12-16 yrs; n = 13) | Adults<br>(17-32 yrs; n = 11) | F/X<sup>2</sup> | p |
| --- | --- | --- | --- | --- | --- | --- | --- | --- |
| **General** |  |  |  |  |  |  |  |  |
| Age at Enrollment | 44 | 12.2 (7.1) | 4.4 (1.3) | 7.5 (1.5) | 14 (1.6) | 21 (4.1) | 106 | **<.001** |
| Age at CS Diagnosis, m (sd) | 44 | 8.0 (6.6) | 3.4 (1.5) | 4.0 (2.2) | 7.4 (3.6) | 16.5 (7.0) | 22.3 | **<.001** |
| Able to Walk Currently (%) | 41 | 34 (83) | 11 (100) | 7 (78) | 9 (82) | 7 (70) | 3.6 | .305 |
| Phrase Speech | 46 | 0% | 0% | 0% | 0% | 0% | - | - |
| **Neurological, n (%)** |  |  |  |  |  |  |  |  |
| *Intellectual Disability* | 42 | 42 (100) | 11 (100) | 9 (100) | 12 (100) | 10 (100) | - | - |
| *Epilepsy* | 42 | 42 (100) | 11 (100) | 9 (100) | 12 (100) | 10 (100) | - | - |
| *Microcephaly* | 41 | 39 (95) | 8 (80) | 9 (100) | 12 (100) | 10 (100) | 6.5 | .089 |
| *Ataxia* | 39 | 37 (95) | 9 (82) | 8 (100) | 11 (100) | 9 (100) | 5.4 | .147 |
| Autism Spectrum Disorder | 40 | 7 (18) | 1 (9) | 2 (29) | 3 (25) | 1 (10) | 2.0 | .574 |
| *Hyperkinesia* | 43 | 40 (93) | 11 (100) | 9 (100) | 11 (92) | 9 (82) | 3.7 | .300 |
| Prior Angelman Dx | 42 | 12 (29) | 0 (0) | 4 (44) | 3 (25) | 5 (50) | 7.8 | .050+ |
| *High Pain Tolerance* | 43 | 39 (91) | 8 (73) | 9 (100) | 12 (100) | 10 (91) | 6.37 | .095 |
| Unprovoked Laughter | 42 | 26 (62) | 5 (45) | 4 (44) | 9 (75) | 8 (80) | 4.69 | .196 |
| Contractures | 39 | 14 (36) | 2 (20) | 3 (33) | 4 (36) | 5 (56) | 2.6 | .451 |
| Visual Acuity Problems | 42 | 22 (52) | 7 (64) | 4 (44) | 6 (50) | 5 (50) | .84 | .841 |
| Eye Movement Abnormalities | 38 | 26 (68) | 6 (60) | 7 (78) | 10 (83) | 3 (43) | 4.1 | .257 |
| **Medical Status, n (%)** |  |  |  |  |  |  |  |  |
| Swallowing Problems | 43 | 19 (44) | 5 (45) | 2 (22) | 5 (42) | 7 (64) | 3.5 | .323 |
| GERD | 41 | 23 (56) | 7 (64) | 2 (22) | 7 (58) | 7 (78) | 6.2 | .103 |
| Current Constipation | 40 | 22 (55) | 6 (55) | 3 (33) | 2 (22) | 11 (100) | 14.6 | **.002** |
| Osteopenia | 39 | 2 (5) | 0 (0) | 0 (0) | 1 (10) | 1 (10) | 2.0 | .572 |
| Current Sleep Problems | 43 | 22 (51) | 7 (64) | 5 (56) | 5 (42) | 5 (45) | 1.3 | .722 |
| Frequent Infections | 41 | 9 (22) | 2 (18) | 2 (22) | 0 (0) | 5 (50) | 7.78 | .051+ |
| # of Hospitalizations, m (sd) | 35 | 4.4 (4.2) | 3.1 (1.4) | 3.3 (3.3) | 3.3 (2.6) | 7.7 (6.2) | 3.01 | **.045** |
| **Regression** |  |  |  |  |  |  |  |  |
| Any Regression, n (%) | 41 | 22 (54) | 3 (30) | 4 (44) | 6 (50) | 9 (90) | 7.9 | **.047** |
| Age at Regression, m (sd) | 17 | 7.2 (6.1) | 2.1 (2.7) | 1.8 (1.6) | 9.2 (3.2) | 11 (7.2) | 3.76 | **.038** |
| Type of Regression, n (%) |  |  |  |  |  |  |  |  |
| Social Skills | 39 | 4 (10) | 0 (0) | 3 (33) | 0 (0) | 1 (11) | 7.6 | .055+ |
| Eating | 40 | 11 (28) | 2 (20) | 2 (22) | 3 (30) | 4 (36) | .87 | .832 |
| Speech | 40 | 9 (23) | 0 (0) | 3 (33) | 1 (9) | 5 (45) | 7.7 | .053+ |
| Motor | 41 | 13 (32) | 1 (10) | 3 (33) | 3 (25) | 6 (60) | 6.1 | .105 |
| **Adaptive Functioning and Behavior** |  |  |  |  |  |  |  |  |
| Irritability (ABC) (sd) | 31 | 4.7 (6.6) | 3.3 (4.7) | 9.2 (12) | 4.8 (6.1) | 3.6 (5.0) | .972 | .420 |
| Hyperactivity (ABC) (sd) | 27 | 22 (15) | 24 (13) | 30 (11) | 23 (17) | 12 (12) | 1.89 | .159 |
| Adaptive Functioning (VABS-2) (sd) | 31 | 35 (13) | 49 (10) | 46 (6.9) | 28 (2.6) | 24 (4.2) | 30.82 | **<.0005** |
| Communication (VABS-2) (sd) | 31 | 34 (10.6) | 46 (11) | 42 (5) | 30 (1.2) | 25 (3.2) | 22.2 | **<.001** |
| Social (VABS-2) (sd) | 31 | 43 (14) | 58 (10) | 53 (5.8) | 36 (4.5) | 31 (11) | 18.5 | **<.001** |
| Daily Living (VABS-2) (sd) | 31 | 35 (12) | 46 (11) | 45 (7.5) | 29 (3.4) | 24 (2.5) | 22.7 | **<.001** |

CS: Christianson syndrome
GERD: gastroesophageal reflux disease
Values with + indicates numbers approaching statistical significance.
VABS-2: Vineland Adaptive Behavior Scales-Second Edition; the Adaptive Function score is a composite of the communication, daily living skills, socialization, and motor skills domain scores
ABC: Aberrant Behavior Checklist, a parent questionnaire to assess behavioral problems in individuals with developmental disorders
Domains presented in italics represent proposed core CS symptoms by age group

Based on our prior study of 14 CS patients, we proposed CS core symptoms (present >85%) and secondary symptoms (present in >35%).^1^ Core symptoms included intellectual disability, non-verbal status, epilepsy, ataxia, postnatal microcephaly, and hyperkinesis.^1^ The previously proposed core symptoms were upheld here in this larger study on average across age by combining all age groups (Table 2). Specifically, all of the core symptoms were present in greater than 85%, including intellectual disability (100%), epilepsy (100%), non-verbal/non-phrase speech (100%), microcephaly (95%), ataxia (95%), and hyperkinesia (93%). Notably, we discerned that high pain tolerance represents a new core symptom when considering all age groups together, present in over 85% (91%).

Secondary symptoms that occurred in 35-84% of participants included current sleep problems (51%), unprovoked laughter (62%), visual acuity problems (52%), current GERD (56%), constipation (55%), eye movement abnormalities (68%), regression (54%), swallowing problems (44%), and contractures (36%) (Table 2). Previously identified secondary symptoms such as prior Angelman syndrome diagnosis and Autism Spectrum Disorder (ASD) diagnosis occurred in 29% and 18% of the current sample, respectively. ASD diagnosis and prior Angelman diagnosis (approaching significance, p = .05) were more common in the older groups, perhaps reflecting more access to accurate genetic testing in the younger group. Scoliosis occurred in 39% of participants; two participants had corrective surgeries for scoliosis between 15 to 20 years of age.

While the core symptoms were relatively stable across age groups, there were several notable exceptions. Absence of phrase speech, intellectual disability, ataxia and epilepsy were stable across all age groups. We found 80% of the youngest age group (below our cut-off for core symptoms) had microcephaly which elevated to 100% in all older age groups, reflecting that idea that the microcephaly in CS has a strong postnatal component (Table 3). This variation in head circumference data reflecting decreases across ages approached significance (p = .089). Similarly, 73% of participants had a high pain threshold in the youngest age group (0-5 years old) which elevated to above 85% (our cut-off for core symptoms) in the older age groups; therefore, although prevalent in the youngest age group, high pain threshold should not be considered a strict core symptom in the youngest age group (Table 3). In the current group, ataxia was present in 82% of participants in the youngest age group, and in 100% of subsequent age groups, again suggesting that ataxia worsens with age. In line with these observations, with regard to the ability to walk, 100% of participants report walking unaided by or before age 5; however, ability to walk unaided is lost in approximately 20% of participants in the 6 to 16 groups and 30% are reported as not able to walk unaided in the oldest group (17+ years old). Finally, with regard to hyperkinesis, this core symptom was present in 100% of the participants in the two younger age groups, and 92% and 82% of the oldest age group, indicating that hyperkinesis does not meet the cut-off for a core-symptom in the oldest age group. Concurrent with these data, we observe that the hyperactivity subscale scores on the Aberrant Behavior Checklist (ABC) are most severe in the younger groups and less prominent in the oldest group (Table 2), although this does not reach statistical significance.

**Table 3.** Core symptoms of Christianson syndrome by age group. List of core symptoms found in >85% of CS males across the following age groups: toddler (2.5-5 years), childhood (6-11 years), adolescence (12-16 years) and adulthood (17-32 years).

| **Core Symptom**<br>*(>85%)* | **Toddlers**<br>*(2.5-5 years)* | **Children**<br>*(6-11 years)* | **Adolescents**<br>*(12-16 years)* | **Adults**<br>*(17-32 years)* |
| --- | --- | --- | --- | --- |
| *Nonverbal* | ✓ | ✓ | ✓ | ✓ |
| *Intellectual Disability* | ✓ | ✓ | ✓ | ✓ |
| *Epilepsy* | ✓ | ✓ | ✓ | ✓ |
| *Microcephaly* |  | ✓ | ✓ | ✓ |
| *Ataxia* |  | ✓ | ✓ | ✓ |
| *Hyperkinesia* | ✓ | ✓ | ✓ |  |
| *High Pain Tolerance* |  | ✓ | ✓ | ✓ |

There were statistically significant differences across developmental groups for some reported symptoms or phenotypic features that are particularly notable. There were significant group differences in the rate of current constipation, prior history of regression, adaptive function (including social, communication, and daily living subscores) and number of hospitalizations (p < .05): each of these experiences were most common in the adult group (Table 2). Importantly, 100% of participants in the oldest age group reported notable problems with constipation (p = .002). With regard to regression, reports of any regression at some point in their lifetime increased significantly with age from toddlers (30%), children (44%), adolescents (50%), and adults (90%) (p = .047). With regard to adaptive functioning, age-normed Vineland scores appeared stable in toddlers and children, and then dropped in the adolescent group and again in the adult group (p < .001 for communication, p < .001 social and daily living, and p < .0005 for adaptive functioning). While not representing a statistically significant difference across groups (p = .305), it is notable that the highest level of inability to walk is experienced in the adult group at approximately 30% (Table 2).

### Longitudinal Assessment of Growth and Physical Stature across the Lifespan in CS

Adult patients with CS have been described as having a thin body habitus^3^; however, systematic analysis of growth in CS across the lifespan has not been conducted. To study growth in CS across the lifespan, raw measurements of height and weight were converted to age-normed percentiles (WHO growth standards for 0-2 years^18^; CDC growth curves for 2-20 years^19^) and to z-scores. To model physical stature across development, we used linear mixed model (with random slopes and intercepts) for each variable of interest (i.e., height, weight) with the age of the child as the time basis. The intercept and age at time point were modeled as fixed and random effects. The height (n = 24) and weight (n = 30) obtained at clinical appointments was analyzed via mixed linear modeling of age-based percentiles (Figure 2). While height and weight naturally increased with time (Figure 2A and 2B), the age-normalized weight, i.e. the z-score (n = 30; total data points = 116; range = 1-12 points per person) linearly declined over time (slope p < .001) (Figure 2C). Height z-score (n = 24; total data points = 81; range = 1-11 points per person) also linearly declined over time (slope p < .001). Of note, the predicted intercept for the model at birth were within normal ranges; however, as shown the model demonstrates a progressive decline in growth in CS relative to normal growth with age (Figure 2). Of relevance to challenges in growth and nutrition, approximately 30% (13/44) participants had G-tubes placed. Approximately 38%, 23%, 23% and 15% had their G-tubes placed during ages of 0-5, 6-11, 12-16, or 17+ years of age. Indications for G-tube placement have been failure to thrive (FTT), most often at the younger ages, and inability to eat across all ages, generally related to challenges with swallowing.

**Figure 2.**
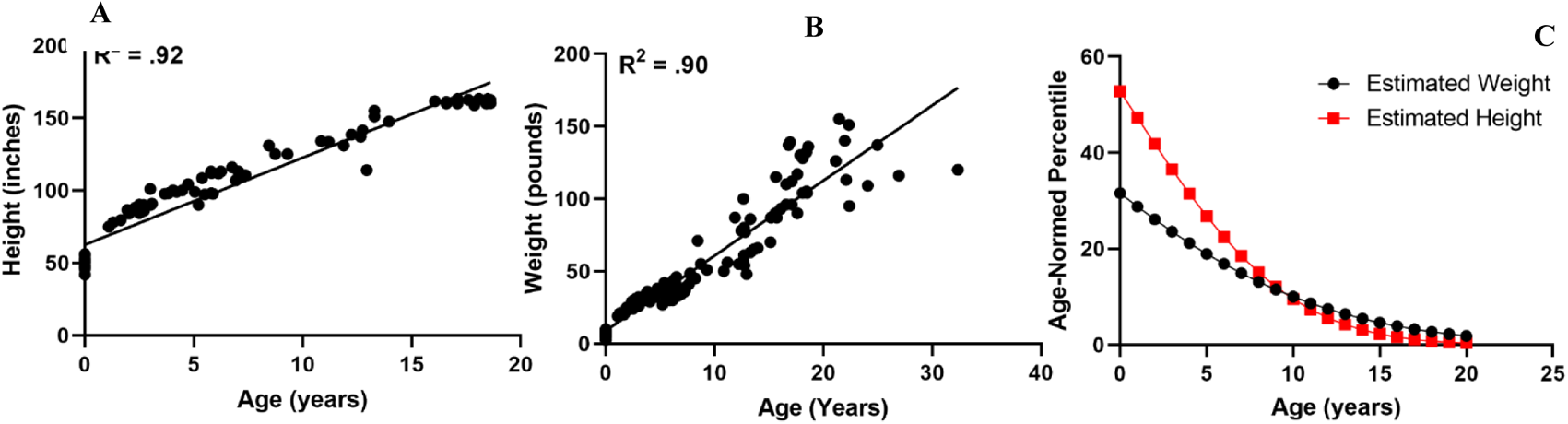
Changes in height and weight across development. (A) Raw score of height (inches) by age (years) from 24 CS probands spanning <1-19 years. Height increases over time (Pearson’s correlation, R^2^=0.92). (B) Raw score of weight (pounds) by age (years) from 30 CS probands spanning <1-19 years. Weight increases over time (Pearson’s correlation, R^2^=0.90). (C) Estimated age-normed height and weight based on liner mixed model analysis. Both height (slope p<0.001) and weight (slope p<0.001) significantly decline over time. Raw height (n = 24, total data points = 81) and weight (n = 30, total data points = 116) measurements were converted to age-normed percentiles (WHO growth standards for 0-2 years^18^; CDC growth curves for 2-20 years^19^) and z-scores. Our linear mixed model (random slopes and intercepts) of age-based percentiles for height and weight modeled the intercept and age at time point as fixed and random effects, respectively. Covariance parameters for height were large relative to their standard errors with statistically significant intercept (slope p=0.003), slope (p=0.016) and covariance of intercept and slope (p=0.046). Covariance parameters for weight were large relative to their standard errors with statistically significant intercept (slope p=0.002), slope (p=0.016) and covariance of intercept and slope (p=0.023).

### Adaptive, Behavioral, and Motor Functioning across the Lifespan

#### Cross-Sectional Findings

Parent-reported adaptive and behavioral data were also available for a subsample of the males (n = 27-31). When examining differences across the cohort by current developmental stage (cross-sectional analysis), a statistically significant difference across groups was identified (p < .0005) in age-normed overall adaptive functioning. Specifically, older participants had lower age-normed abilities than younger participants, in that the younger participants had abilities more similar to their same-aged peers while the abilities of the older participants were further below those of their same-aged peers (Table 2). In further examining across these cross-sectional age groups, there were no significant differences between age groups in hyperactivity or irritability at baseline (Table 2; ABC).

#### Longitudinal Adaptive/Motor Findings

Importantly, we also conducted a longitudinal follow-up analysis through investigation of change in adaptive function in individuals (n = 22), at baseline and at a 1-year follow-up time point. The change in skills between baseline and follow-up was correlated to age at enrollment. Results showed that participant age was negatively correlated with change in fine motor skills (r = −.50; p = .028) and gross motor skills (r = −.63; p = .002; Figure 3). Specifically, 5 of 6 adults (83%) experienced a loss in multiple fine motor skills such as picks up small objects with thumb and fingers and moves object from one hand to the other. Expressive language and interpersonal skills change were not significantly correlated with age (Figure 3). These longitudinal studies suggest that older participants are at risk for losing previously acquired skills, particularly in the motor domain, which is observable by parents/guardians at a one-year follow-up. Therefore, these longitudinal studies augment the above cross-sectional studies.

**Figure 3.**
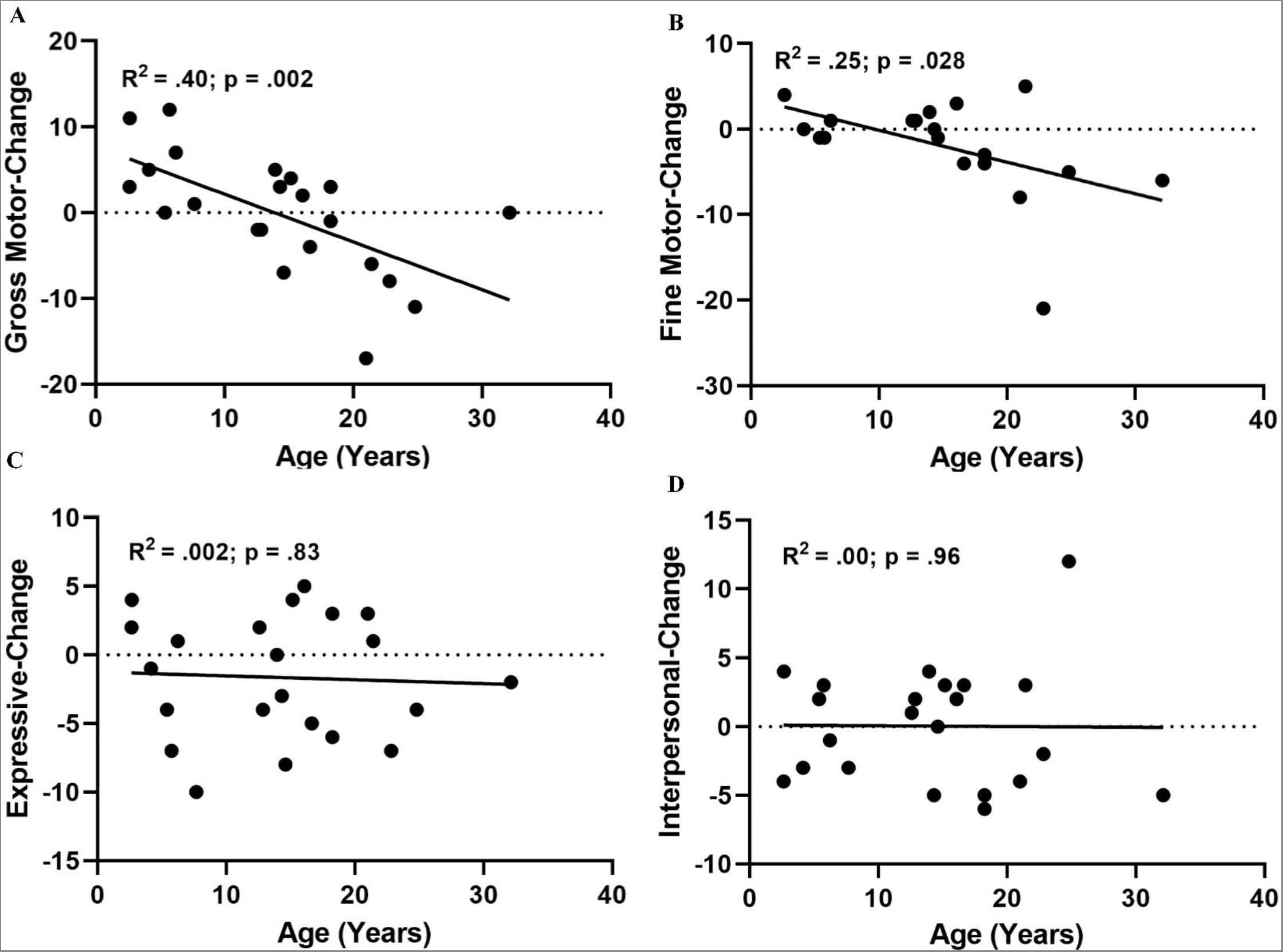
Longitudinal changes in motor, language, and social functioning over 1-year period. Parent-rated changes in gross motor (**A**), fine motor (**B**), expressive language (**C**), and interpersonal/social functioning (**D**) from enrollment to 1-year follow-up (n = 22). Each point represents an individual proband. Raw score change between enrollment and 1-year follow-up is plotted by age at enrollment. Changes in gross (**A**, R^2^=0.40, p=0.002) and fine (**B**, R^2^=0.25, p=0.028) motor is significantly correlated with age, as older age is associated with greater loss of motor skills over a 1-year period. There is no association between age and changes in expressive language (**C**, R^2^=0.002, p=0.83) or social functioning (**D**, R^2^=0.00, p=0.96) over this timespan. Pearson’s correlation.

#### Neurological Exam

Standardized, in-person motor examination was performed by a board-certified neurologist (JSL). Twelve patients were directly examined (Supplemental Table 2). The direct assessment of patients additionally supports the interpretation that there is age-related worsening of motor function. Older patients (i.e. age 20 and greater), present with evidence of motor dysfunction with signs of corticospinal tract, i.e. upper motor neuron damage, such as increased tone, weakness, increased and abnormal reflexes. These older subjects also have evidence of cerebellar dysfunction including tremor and truncal ataxia, as evidenced by wide based gait. Thus, the clinical phenotype includes upper-motor neuron involvement that emerges particularly in adulthood, as well as cerebellar dysfunction that is also progressive but presents first in early adulthood.

### Participant Deaths

Three of the 44 participants in this described cohort died (6.8%) during the duration of the study. One participant, who also may have had other contributing genetic mutations, died between 6 to 10 years of age due to complications caused by a worsening neurological state and increased seizure frequency. The adult participants died between the ages of 20 to 25 and 30 to 35 years in their homes due to conditions related to infirmity, such as pneumonia.

## DISCUSSION

This study represents the largest to date of patients with Christianson Syndrome (CS) with confirmed NHE6 mutations. This also is the first study to date as part of the International Christianson Syndrome and NHE6 (SLC9A6) Gene Network Study, presenting longitudinal follow-up and systematic study of CS into adulthood. Forty-four individuals with 31 unique NHE6 mutations, age 2 to 32 years, were followed prospectively, herein reporting baseline, 1-year follow-up, and retrospective natural history. Among the most prominent results are: first, the large range of mutations, wherein there are some notable new missense mutations (discussed below); second, we have also defined the core symptoms of CS and the dynamics of these symptoms across the lifespan, and also, added a novel and important new core symptom, namely the high pain threshold that children exhibit, particularly after age 6; third, we present longitudinal follow-up data demonstrating that CS patients do show decreases in motor function with age; fourth, we present data exhibiting the challenges in maintenance of a healthy weight and reductions of body-mass index with age; and finally, fifth, in this study through longitudinal follow-up, we were able to provide important information with regard to CS features in adulthood and we begin to address the question of life expectancy and mortality. This international CS study has thereby defined future treatment targets, and also provides natural history data of the sort that will support future clinical trials.

### Genetic Diversity in CS and Genotype-Phenotype Relationships

We present 31 unique mutations in 44 affected males with CS. The vast majority are clear complete or near complete loss-of-function mutations, with more than 80% as frameshift/nonsense, splice (29% of total) or highly deleterious copy number variant deletions. A minority represent missense mutations which are likely loss-of-function. There is a possibility that these missense mutations may represent incomplete loss-of-function mutations; however, at present the evidence to support this is only anecdotal. In the current dataset, there is no statistical evidence of genotype-phenotype correlations, providing further support that the majority of *NHE6* mutations are complete or near complete loss-of-function. Prior studies in patient-derived induced pluripotent stem cells have demonstrated that nonsense mutations are subject to nonsense mediated mRNA decay.^14^ Of note, the G383D mutation which has been studied functionally in iPSCs exhibits some residual proteins.^14^ Another notable finding, in terms of genetic mutations, is that we present here a patient with the L582P mutation which represents a rare missense mutation in the cytoplasmic tail of NHE6 that is associated with the CS phenotype. The proband with this L582P mutation exhibits core CS symptoms including intellectual disability, non-verbal, and microcephaly.

### Natural History of Primary and Secondary CS Symptoms with Age

Previously established core diagnostic features of CS^1^ were confirmed in this sample of probands, including intellectual disability (100%), epilepsy (100%), non-verbal/non-phrase speech (100%), postnatal microcephaly (95%), ataxia (95%), and hyperkinesia (93%). High pain tolerance, not previously identified as a core feature of CS, was present in 91% of probands, and therefore, we support the inclusion of high pain tolerance as a new core diagnostic feature. Importantly, clinicians and caregivers should be aware of this symptom, in order to understand how people with CS may respond to injury. Minimal response to pain may not be a good indicator in the evaluation of the seriousness of an injury. Overall, with regard to core symptoms of CS, we show here that there is some evolution of these symptoms with age, such that microcephaly, ataxia and high pain threshold are not present in over 85% of CS patients until age 6. Notably, this provides further support that the microcephaly in CS is indeed postnatal microcephaly, and generally not primary microcephaly. Furthermore, hyperkinesia appears to dissipate with age, which may be mediated by age-related worsening of motor function.

Secondary symptoms of CS^1^ were also highly represented in the sample, including GERD (56%), eye movement abnormalities (68%), prior regression (54%), and prior diagnosis of Angelman syndrome (29%) and autism (18%). The majority of these symptoms were similarly observed across developmental periods. However, we suspect a prior diagnosis of Angelman syndrome will become less common in younger CS males in the future due to the availability and advances in genetic testing. Indeed, no toddlers were previously diagnosed with Angelman syndrome. Additional symptoms such as current sleep problems (51%), visual acuity problems (52%), and swallowing problems (44%) were relatively stable across developmental periods. Importantly, the adult cohort experienced a higher rate of select medical complications, particularly gastrointestinal symptoms, such as gastroesophageal reflux, and constipation. Adult CS patients had significantly greater hospitalizations, along with more frequent infections. Unprovoked laughter becomes more prominent in adolescence/adulthood, whereas in Angelman syndrome laughter appears to decrease with age.^20–22^ Also, importantly, scoliosis occurred in greater than a third of patients which worsens with age, requiring surgery in a subset.

### Challenges in Maintaining a Healthy Weight During Growth and Aging in CS

Clinical records were analyzed to examine height and weight growth across development as they represent secondary CS symptoms.^1^ While probands continued to make raw score gains in pounds and inches across time, their slowed rate of growth resulted in a similar pattern of decreasing age-normed score across development for height and weight. Mixed model results estimated height and weight to be in the average or typically developing range at birth. However, height fell to below one standard deviation (SD) below the mean by 7 years and two SDs by 19 years old. Weight fell to below one SD by 8 years old and two SDs by 15 years old. While there was little evidence of loss in body mass in our data, models were only calculated from 0-20 years (based on available developmental normative data). These analyses of body size highlight abnormalities in growth in CS; however, these data call for important future studies. Healthy weight should become a target symptom in growth and adulthood. Studies of nutrition and healthy weight in adulthood are warranted, and consideration of the caloric and nutritional requirements of men with CS will need to become a critical subject in future studies.

### Longitudinal Data in CS Demonstrate Progression of Motor Symptoms

We present natural history data reflecting both retrospective methods, as well as prospective study at 1-year follow-up and cross-sectional information on families. The majority of probands (54%) had a history of parent-reported developmental regression that increased in prevalence across development, with 30% of toddlers, 44% of children, 50% of adolescents, and 90% of adults experiencing prior regression. This was not specific to any of the subdomains, although 60% of adults had a prior motor regression and 45% of adults had a prior speech regression. This is consistent with our prior study of regression in 50% of CS probands, most notably in speech and motor domains.^1^ Additionally, loss of social skills was noted in about one-third of children with regressions.

To complement these and prior cross-sectional and retrospective analyses, we examined changes in language, social, and motor functions over a 1-year interval follow-up. Here, we find that there was a negative association between participant age at enrollment and fine/gross motor function loss, in that adults experienced a notable loss of motor functions in this 1-year interval. These data identify motor function as a target symptom for treatment and prevention of decline. These prospective follow-up data are particularly notable as we are able to observe a measurable decline in this relatively short time-line of 1-year. This finding is particularly valuable and hopeful as clinical trials that may target the progression of these motor symptoms will benefit from observations of this strong effect size in time-frames that appear feasible for a robust trial.

### Limitations of this study

While this is the largest study to date on CS, the sample size (n = 44) is modest, and definition of more rare events in CS will require a larger and more highly-powered sample. Additionally, while this study does include prospective, intra-individual longitudinal data, many findings still rely on natural history data from retrospective clinical records or a convenience sample. For example, this study relies on a clinical diagnosis of autism, as opposed to prospectively measuring autism symptoms using standardized assessments such as the Autism Diagnostic Observation Schedule (ADOS)^23^, Autism Diagnostic Interview-Revised (ADI-R)^24^, and Social Communication Questionnaire (SCQ)^25^. Fewer CS patients were clinically diagnosed with autism (18%, 7 out of 40) compared to our prior study (43%, 6 out of 14).^1^ While there is a strong agreement between a clinical diagnosis and these measurements^26–28^, we may be underestimating autism-related features in CS. Indeed, in our prior study using standardized assessments in a small subset of CS patients, we found a majority met diagnostic criteria for autism by the ADI-R (100%, 3 out of 3) and SCQ (89%, 8 out of 9).^1^ In addition, this study does not examine the clinical progression of epilepsy symptoms, treatment and outcome. Our prior study had a detailed cross-sectional analysis^1^, and an in-depth analysis of epilepsy natural history is beyond the scope of the current manuscript and is being addressed in a parallel study. Of note, Dr. Arnold Christianson in his original study in a large South African pedigree suggested that there may be a risk of premature mortality in CS. In the course of our study, we have observed the death of three total participants. While this may appear as an elevated risk given this profound neurologic condition, our sample size remains too small to discern the specific underlying susceptibilities that may put males with CS at an increased mortality risk in childhood or in aging. Again, it currently remains unclear whether premature mortality is a risk factor in CS.

### Summary

As a part of the International Christianson Syndrome and NHE6 (SLC9A6) Gene Network study, in collaboration with the Christianson Syndrome Association, we present the largest study to date of male patients with CS. Overall, this study has identified a large range of loss-of-function mutations, including several novel missense variants. This study is notable for the identification of a new core symptom (high pain threshold) and the natural progression into adulthood. This study has the largest group of adults with CS, and we have defined the clinical progression of motor symptoms. In addition, we identify a decline in body-mass index with age as an important treatment target moving forward. In conclusion, this study identifies several target symptoms – particularly health weight gain and motor symptoms -- that progress with a natural history that may make them amenable to intervention in a future clinical trial. As this CS study grows, the findings presented here, and in future studies, will enhance clinical readiness as targeted treatments become available.

## Data Availability

All data produced in the present study are available upon reasonable request to the authors.

## Acknowledgements

The authors would like to thank the families for participating in this study. We would also like to thank the Christianson Syndrome Association for their help with this study. This work was supported by National Institutes of Health Grants R01NS113141, R01MH102418, R01MH105442, and R21MH115392 to E.M.M. EMM had full access to all of the data in the study and takes responsibility for the integrity of the data and the accuracy of the data analysis. None of the authors have a financial conflict of interest.

## SUPPLEMENTARY MATERIALS

**Supplementary Table 1.** Clinical characteristics of CS males by inheritance and mutation type. Adaptive functioning score from VABS-2 is a composite of the communication, daily living skills, socialization and motor skills domain scores. GERD=Gastroesophageal reflux disease. ABC=Aberrant Behavior Checklist. VABS-2=Vineland Adaptive Behavior Scales-2^nd^ Edition.

|  | De Novo<br>(n = 15) | Inherited<br>(n = 22) | *t*/*X*<sup>2</sup><br>*p* | Missense<br>(n = 6) | Nonsense<br>(n = 22) | *t*/*X*<sup>2</sup><br>*p* |
| --- | --- | --- | --- | --- | --- | --- |
| **General** |  |  |  |  |  |  |
| Age at Enrollment (sd) | 12 (5.9) | 13 (7.8) | .603 | 8.1 (6.6) | 14 (5.8) | **.041** |
| Age at CS Diagnosis, m (sd) | 7.1 (5.7) | 8.6 (7.6) | .523 | 6.1 (4.5) | 8.5 (6.7) | .403 |
| Able to Walk Currently (%) | 11 (85) | 17 (81) | .785 | 5 (83) | 16 (76) | .959 |
| History of Regression (%) | 7 (54) | 11 (52) | .934 | 2 (40) | 11 (52) | .619 |
| **Neurological, n (%)** |  |  |  |  |  |  |
| Microcephaly | 14 (100) | 19 (95) | .396 | 5 (100) | 20 (100) | - |
| Ataxia | 14 (100) | 17 (94) | .370 | 6 (100) | 17 (94) | .555 |
| Autism Spectrum Disorder | 4 (29) | 2 (10) | .162 | 1 (17) | 5 (25) | .671 |
| Hyperkinesia | 14 (100) | 19 (86) | .149 | 3 (100) | 18 (75) | .326 |
| Prior Angelman Dx | 4 (29) | 8 (38) | .561 | 2 (33) | 7 (35) | .940 |
| High Pain Tolerance | 14 (100) | 19 (86) | .149 | 6 (100) | 19 (90) | .432 |
| Unprovoked Laughter | 6 (43) | 16 (76) | **.046** | 4 (67) | 14 (70) | .877 |
| Contractures | 5 (36) | 5 (28) | .631 | 2 (40) | 5 (28) | .599 |
| Visual Acuity Problems | 9 (64) | 9 (43) | .214 | 4 (67) | 10 (50) | .473 |
| Eye Movement Abnormalities | 9 (69) | 14 (70) | .963 | 3 (60) | 14 (74) | .549 |
| **Medical Status, n (%)** |  |  |  |  |  |  |
| Swallowing Problems | 7 (100) | 8 (36) | .418 | 3 (50) | 8 (38) | .601 |
| GERD | 8 (57) | 11 (52) | .782 | 3 (50) | 11 (55) | .829 |
| Current Constipation | 5 (42) | 13 (62) | .261 | 4 (67) | 11 (61) | .808 |
| Osteopenia | 2 (14) | 0 (0) | .089 | 1 (17) | 1 (6) | .394 |
| Current Sleep Problems | 6 (43) | 12 (55) | .494 | 3 (50) | 11 (52) | .918 |
| Frequent Infections | 3 (23) | 5 (24) | .961 | 3 (50) | 3 (16) | .087 |
| # of Hospitalizations, m (sd) | 3.9 (2.7) | 5.0 (5.4) | .521 | 3.2 (1.5) | 4.4 (3.8) | .570 |
| **Adaptive Functioning and Behavior** |  |  |  |  |  |  |
| VABS-2: Adaptive Function | 35 (13) | 34 (14) | .839 | 40 (12.5) | 30 (9.4) | .082 |
| ABC: Irritability | 6.0 (6.3) | 4.9 (7.4) | .704 | 5 (7.8) | 4.6 (5.6) | .920 |
| ABC: Hyperactivity | 31 (6.0) | 21 (16) | .136 | 35 (11.8) | 21.6 (13.6) | .135 |

VABS-2: Vineland Adaptive Behavior Scales-Second Edition; the Adaptive Function score is a composite of the communication, daily living skills, socialization, and motor skills domain scores
ABC: Aberrant Behavior Checklist, a parent questionnaire to assess behavioral problems in individuals with developmental disorders
GERD: gastroesophageal reflux disease

**Supplementary Table 2.** Neurological examination of CS probands. Neurological examination of 12 CS males. L=Left. R=Right. L>R=Left greater than right. R>L=Right greater than left. s.p.=Status post.

| Age @ Exam | Mutation cDNA position | Mutation Protein Position | Vocalizations | Extraocular Movements | Tone, limb or side | Strength, or pattern of weakness | Tremor | Deep tendon reflex (DTR) | Babinski, left or right | Gait: base, posture |
| --- | --- | --- | --- | --- | --- | --- | --- | --- | --- | --- |
| 6-10 | c.1380_1389delinsG TCTTGG GAAGAC T | p.N460Kfs*12 | Present | Esotropia, L>R | Normal | Full | None | Normal and symmetric | Absent | Wide base |
| 11-15 | c.680+5G>A | - | Present | Esotropia, s.p. surgical correction | Normal | Full | None | Could not assess | Could not assess | Wide base |
| 11-15 | c.899+1del GTAA | - | Present | Esotropia, L | Could not assess | Full | None | Normal and symmetric | B/L | Unable to walk |
| 11-15 | c.680+1G>A | - | ~ 5 words | Normal | Normal | Full | None | Normal and symmetric | Absent | Normal |
| 16-20 | c.526-1>A | - | Present | Strabismus, s.p. surgical correction | Low-normal | Could not assess | None | High amplitude patellar reflexes with crossed-adductor response | Right | Wide base |
| 16-20 | c.630_631insT | p.I211Yfs*53 | Present | Strabismus, s.p. incomplete surgical correction | Low-normal | Full | Upper extremity action tremor | High amplitude throughout | Left | Wide base |
| 21-25 | c.540_547dup | p.F183fs*1 | Present | Normal | Increased tone, R>L | Full | Left arm intermittent postural tremor | Normal and symmetric | Absent | Wide base |
| 21-25 | 120.7kb deletion from intron 10 through 3'-untranslated region | - | Present | Normal | Increased tone, L>R | Mild Left hemiparesis | None | Normal and symmetric | Left | Posturing of upper extremities |
| 26-30 | c.630_631insT | p.I211Yfs*53 | Present | Normal | Increased tone, Left leg | Full | None | High amplitude throughout; symmetric | B/L | Wide base with toe walking |
| 26-30 | c.1148G>A | p.G383D | Present | Normal | Normal | Full | None | Normal and symmetric | Absent | Wide base |
| 26-30 | c.1569G>A | p.W523* | Present | Normal | Normal | Full | Left leg tremor | Normal and symmetric | Unable to assess | Wide base |
| 26-30 | c.630_631insT | p.I211Yfs*53 | Present | Strabismus, s.p. surgical correction | Increased tone | Full | Upper extremity action tremor | High amplitude throughout | B/L | Wide base, diminished arm swing |

## Notes

### Competing Interest Statement

The authors have declared no competing interest.

### Funding Statement

This study was funded by National Institutes of Health Grants R01NS113141, R01MH102418,
R01MH105442, and R21MH115392 to E.M.M.

### Author Declarations

IRB of Lifespan Healthcare/Brown University gave ethical approval for this work.

